# Fast SARS-CoV-2 detection by RT-qPCR in preheated nasopharyngeal swab samples

**DOI:** 10.1101/2020.04.08.20058495

**Authors:** Julia Alcoba-Florez, Rafaela González-Montelongo, Antonio Íñigo-Campos, Diego García-Martínez de Artola, Helena Gil-Campesino, The Microbiology Technical Support Team, Laura Ciuffreda, Agustín Valenzuela-Fernández, Carlos Flores

**Affiliations:** Servicio de Microbiología, Hospital Universitario N. S. de Candelaria, Santa Cruz de Tenerife, Spain; COVID-19 Study Group, Servicio Canario de la Salud, Santa Cruz de Tenerife, Spain; Genomics Division, Instituto Tecnológico y de Energías Renovables, Santa Cruz de Tenerife, Spain; Research Unit, Hospital Universitario N. S. de Candelaria, Santa Cruz de Tenerife, Spain; Laboratorio de Inmunología Celular y Viral, Unidad de Farmacología, Facultad de Medicina, Universidad de La Laguna, San Cristóbal de La Laguna, Spain; Red española de Investigación en VIH/SIDA (RIS)-RETIC, Instituto de Salud Carlos III, Madrid, Spain; CIBER de Enfermedades Respiratorias, Instituto de Salud Carlos III, Madrid, Spain; Instituto de Tecnologías Biomédicas (ITB), Universidad de La Laguna, San Cristóbal de La Laguna, Spain

**Author notes:** M. Sonia Batista-Torres, Concepción Beltrán-Tacoronte, Esther G. Gómez-Ruíz, Victoria González-González, Teodora Manrique-Izquierdo, Rocio M. Rivera-Ruiz, Iru Trujillo-Medina, Guacimara Espinel-Guerra, Chaxiraxi Medina-Coello, M. Candelaria Padilla-Martín. **Correspondence:** Dr. Carlos Flores, Research Unit, Hospital Universitario N. S. de Candelaria, Carretera del Rosario s/n, 38010 Santa Cruz de Tenerife, Spain.

**Keywords:** COVID-19, SARS-CoV-2, diagnosis, sample treatment, RNA extraction, fast protocols

## Abstract

The current reference for COVID-19 diagnosis is based on the detection of SARS-CoV-2 on RNA extracts using one-step retrotranscription and quantitative PCR (RT-qPCR). Based on the urgent need for high-throughput COVID-19 screening, we tested the performance of three alternative, simple and affordable protocols to rapidly detect SARS-CoV-2, overcoming the long and tedious RNA extraction step. Although with an average increase of 6.1 (± 1.6) cycles compared to standard tests with RNA extracts, we show that RT-qPCR yielded consistent results in nasopharyngeal swab samples that were subject to a direct 70°C incubation for 10 min. Our findings provide viable options to overcome any supply chain issue and help to increase the throughput of diagnostic tests by using any qPCR device, thereby complementing standard COVID-19 testing.

## Introduction

The ongoing coronavirus disease 2019 (COVID-19) pandemic due to Severe Acute Respiratory Syndrome Coronavirus 2 (SARS-CoV-2) worldwide infection (https://www.who.int/emergencies/diseases/novel-coronavirus-2019/situation-reports) has imposed an unexpected high burden on the health care systems worldwide leading to an increasing demand for daily diagnostic screening. The current standard assay for diagnosis is based on the extraction of RNA from respiratory samples, especially from nasopharyngeal swab viral transport media (VTM), and subsequent one-step reverse transcription and real-time quantitative PCR (RT-qPCR) targeting one or several sequences from SARS-CoV-2 (Corman *et al*., 2020). However, this standard procedure usually takes 3.5-4.0 h considering the manual interventions and there is a risk of reagent shortage in major kit suppliers, particularly for the RNA extraction step. Besides, the common hospital practice relies on liquid-handling robots for RNA extractions that adds another bottleneck nowadays given the large number of populations being screened.

Here we aimed to simplify the current diagnostic standard for COVID-19 by overcoming the RNA extraction step. We tested three simple approaches based on direct nasopharyngeal swab VTM heating before the RT-qPCR: a) directly without additives (Direct); b) in a formamide-EDTA (FAE) buffer (Shedlovskiy, Shcherbik and Pestov, 2017); and c) in a RNAsnap™ buffer (Stead *et al*., 2012). As a reference, we compared their results with those obtained using a standard RNA extraction protocol on the same samples.

## Materials and Methods

### Ethics statement

The study was conducted at the University Hospital Nuestra Señora de Candelaria (Santa Cruz de Tenerife, Spain) during March 2020. The institutional review board approved the study (ethics approval number: CHUNSC_2020_24).

### Exploratory stage

#### Samples

We selected nasopharyngeal swabs from four COVID-19/SARS-CoV-2 patients and four COVID-19 negative controls collected in 2 mL volume of VTM (Deltalab). Sample manipulation was performed under a biosafety class II cabinet (TELSTAR bio-II-A). Diagnosis of COVID-19/SARS-CoV-2 infection was conducted with 200 μL of VTM with standard RNA extractions, which used the MagCore viral Nucleic Acid Extraction kit (RBC Bioscience) in a MagCore liquid handler (RBC Bioscience) (lasting approximately 1 h) or the Nuclisens easyMAG kit (bioMérieux) with the eMAG liquid handler (bioMérieux) (lasting approximately 1 h and 20 min), and a one-step RT-qPCR using the LightMix^®^ Modular SARS and Wuhan CoV E-gene kit (Roche). This commercial mix was supplemented with primers and probes for detecting the human actin (*ACTB*) gene expression to serve as an internal control of the reaction as described elsewhere (Fenollar *et al*., 2016) (**Table 1**). The experiments included a non-template control and a positive control for the E-gene amplification (LightMix^®^ kit Sarbecovirus ivRNA control +, Roche). The RT-qPCR was performed in 10 μL final volume reactions (5 μL of sample) using a CFX96 Touch Real-Time PCR Detection System (Bio-Rad). Following the specifications, thermal cycling was performed at 55°C for 5 min for the RT step, followed by an amplification step with initial denaturation at 95°C for 5 min and 45 cycles of 95°C for 5 sec, 60°C for 15 sec, and 72°C for 15 sec. A final cooling step at 40°C for 30 sec was also included. Thermal cycling took 1 h and 14 min in total.

**Table 1.**
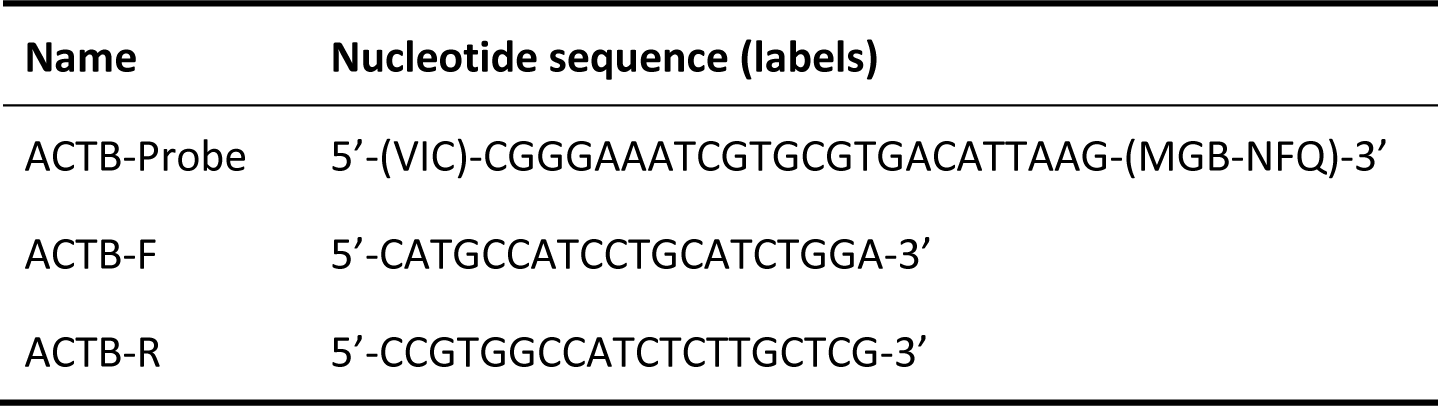
Oligonucleotide sequences used to detect the *ACTB* gene expression as an internal reaction control for SARS-CoV-2 detection.

#### Alternative sample treatments

Fresh FAE and RNAsnap™ buffers were prepared from Molecular Biology grade reagents following the described conditions (Stead *et al*., 2012; Shedlovskiy, Shcherbik and Pestov, 2017). Aliquots of 100 μL from each VTM sample were mixed with 10 μL of each buffer using a vigorous vortexing and heated at 70°C for 10 min. Note that the RNAsnap™ procedure was originally described to be used at 95°C for 7 min. In parallel, a 10 μL aliquot of each sample was also heated at 70°C for 10 min. RT-qPCR was performed with 3.5 μL temperature-treated sample aliquots from each treatment in a 10 μL final volume using the same conditions as described for the standard diagnosis of COVID-19/SARS-CoV-2 infection.

### Validation stage

#### Samples and treatment

We selected nasopharyngeal swab samples from 90 independent donors where diagnosis of COVID-19/SARS-CoV-2 infection was available following the same methods indicated above (41 COVID-19 positives and 49 negatives). The experiments also included a non-template control and a positive control for the E-gene amplification. These samples were subject to the treatment that provided the results with the smallest cycle threshold (Ct) deviations from the standard protocol in the exploratory stage. The experiment was blind to the COVID-19 diagnosis assignment. Then, the resulting confusion matrix was used to obtain sensitivity, specificity and accuracy of the alternative sample treatment compared to the assays based on standard RNA extractions.

## Results

### Exploratory stage

We ensured that the non-template control did not amplify in any of the protocols both for the SARS-CoV-2 or the *ACTB*. Similarly, the positive control for the E-gene amplification yielded positive results in the RT-qPCR experiments of the three alternative sample treatment protocols. The use of specific and highly sensitive probes for coronavirus E-gene amplification leads to optimal results, therefore, the rationale for the first tests design has been followed in our assays (Corman *et al*., 2020). Of note, this envelope small membrane protein (E) is required to produce a structurally complete viral particle together with three other major structural proteins: the spike (S), the nucleocapsid (N) and the membrane (M) proteins (Mortola and Roy 2004; Masters 2006; Wang 2017). All samples gave positive results for the internal human *ACTB*. Ct for the E-gene amplification in the standard RNA extractions indicated that two of the positive COVID-19 cases had low Ct (<20) while the other two had large values (Ct>28) (**Table 2**).

**Table 2.**
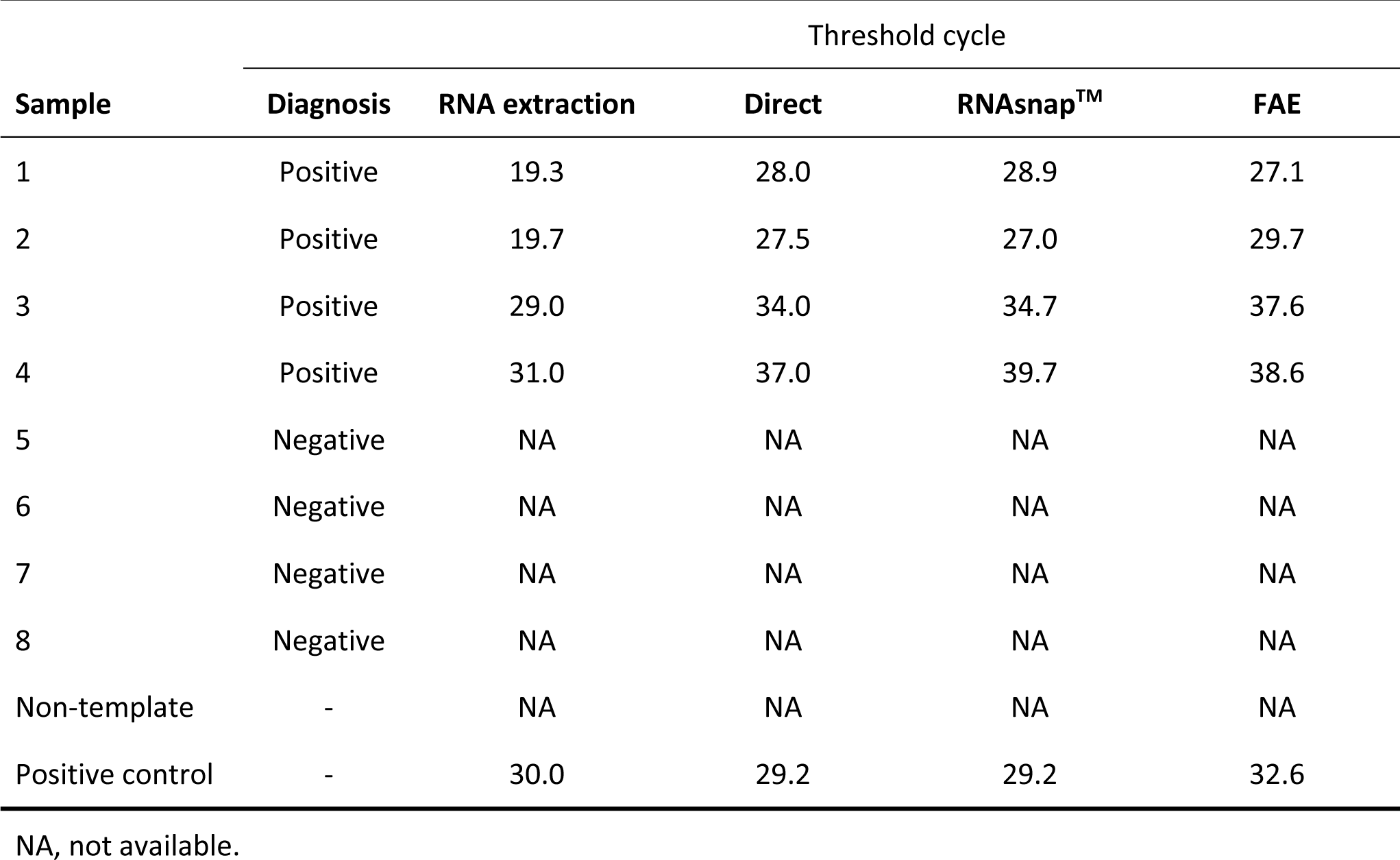
Main RT-qPCR results for SARS-CoV-2 E-gene amplification.

The four COVID-19/SARS-CoV-2 positive cases amplified in the three treatments. None of the COVID-19/SARS-CoV-2 negative samples amplified for the E-gene in these treatments. Although Ct values for the E-gene of the COVID-19/SARS-CoV-2 positive samples were concordant within samples among treatments, we observed a displacement of the Ct in all the positive samples using the three alternative treatments compared to the standard RNA extractions (**Table 2**). Compared to the standard RNA extraction, we observed for the E-gene an average (± SD) increase in the Ct of 6.9 (± 1.7), 7.8 (± 1.7), and 8.5 (± 1.1) for the Direct, the RNAsnap™ and FAE treatments, respectively (**Figure 1**).

**Figure 1.**
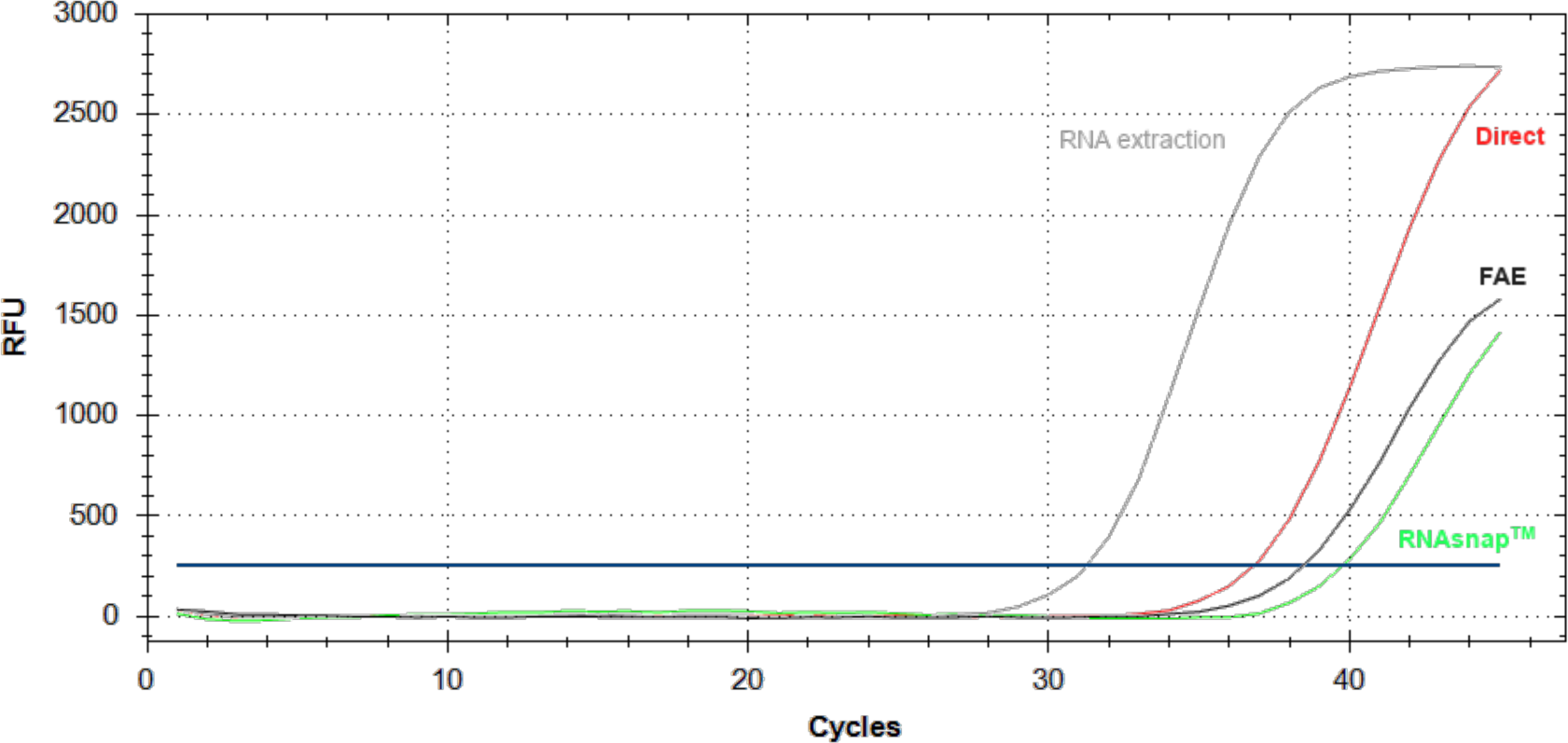
RT-qPCR amplification plots for the SARS-CoV-2 E-gene for a nasopharyngeal swab VTM sample subjected to the standard RNA extraction, and the Direct, the RNAsnap™ and FAE treatments. The horizontal blue line denotes the cycle threshold. RFU: Relative fluorescence units.

### Validation stage

Based on these results, we then assayed 90 VTM samples from independent donors using the Direct method under the same procedures as in the previous stage. In this reaction, the non-template control did not amplify and the positive control for the E-gene amplification yielded a positive result. We verified that all samples gave positive results for the internal human *ACTB* (average Ct of 29.6 ± 2.5) although the amplification Ct was, on average, slightly larger than that obtained by the standard RNA extraction method in the same samples (average Ct of 27.0 ± 1.5).

Out of the 41 COVID-19/SARS-CoV-2 positive VTM samples, only five did not yield amplification for the E-gene with the Direct treatment. Regarding the *ACTB* gene amplification results on the extracted RNA of these five samples, we did not observe significant differences when compared with those from the other COVID-19/SARS-CoV-2 positive samples (average Ct of 27.6 ± 1.2 and 26.9 ± 1.6, respectively; p=0.457). However, their Ct values for the E-gene were significantly larger (average Ct of 34.0 ± 2.0 and 25.7 ± 4.9, respectively; p=0.0007). Therefore, we considered these five samples as false negatives, corresponding to a false negative rate in the Direct treatment of 12% (95% confidence interval [CI]= 5-28). Considering the 36 samples that were COVID-19/SARS-CoV-2 positive by the two methods, there was an average increase in the E-gene Ct by the Direct method of 6.1 (± 1.6) compared to that obtained by a regular RNA extraction. None of the COVID-19/SARS-CoV-2 negative VTM samples was classified as a positive by the Direct treatment. Therefore, the Direct method yielded a sensitivity, specificity and accuracy of 87.8% (95% CI= 73.8-95.9), 100% (95% CI=92.8-100), and 99.9% (95% CI= 95.7-100), respectively.

## Discussion

While the three heating treatments of the sample and direct use in the subsequent detection demonstrated positive amplification of the SARS-CoV-2 E-gene, the Direct method provided the best results and were highly consistent with the COVID-19/SARS-CoV-2 infection diagnosis based on the standard RNA extraction in nearly half of the time (**Figure 2**). We warn that while we observed amplification of other SARS-CoV2 target alternatives of the same kit on samples tested using the Direct method (i.e., the LightMix^®^ Modular SARS and Wuhan CoV N-gene and RdRP kits, Roche) (not shown), they are likely to have a reduced sensitivity as compared to that calculated for the E-gene.

**Figure 2.**
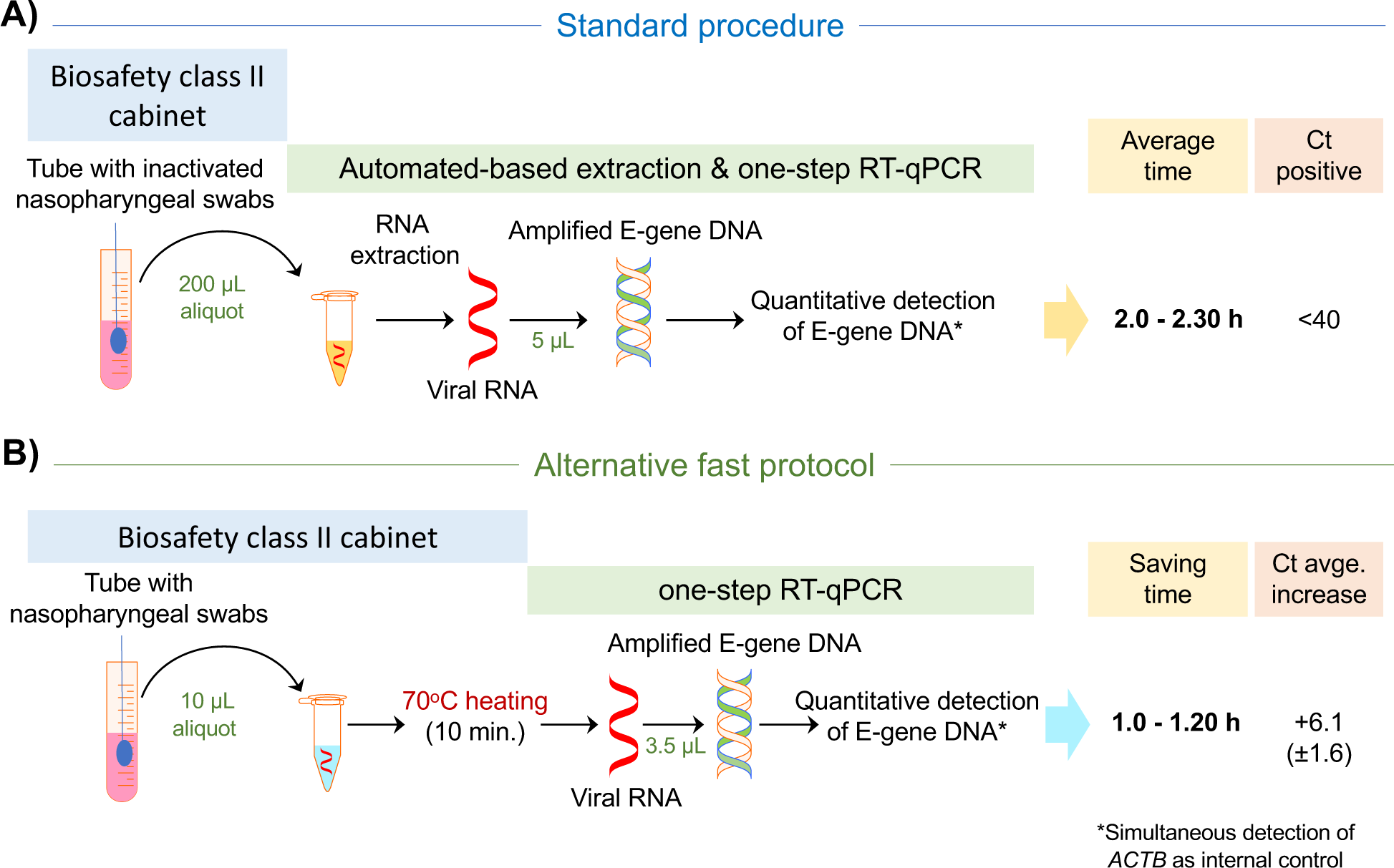
Schematic diagram of the standard RNA extraction-based protocol (A) compared to the alternative fast protocol (B) and the timesaving estimates for RT-qPCR-based SARS-CoV-2 testing. Indicated times should be taken as conservative estimates as they will be dependent on the personnel skills and the number of samples being assessed on the experiment. Ct, cycle threshold.

While the performance of this alternative, simple, and affordable protocol will help to accelerate COVID-19/SARS-CoV-2 infection diagnosis, we caution that the study was done with a limited number of samples and amplifications should be closely monitored to avoid increasing the false negatives above that of the standard diagnosis based on RNA extractions (Xie *et al*., 2020).

Given the comparable environmental stability of SARS-CoV-2 and SARS-CoV-1, the most closely related human coronavirus (van Doremalen *et al*., 2020), and the evidence suggesting that SARS-CoV-1 loses infectivity above 56°C within short periods of time (Geller, Varbanov, and Duval, 2012), we postulate that the Direct protocol may also help to control the infectiveness of the manipulated samples during COVID-19/SARS-CoV-2 infection diagnosis.

We feel these results provide viable options to overcome any supply chain issue, helping to increase the throughput of COVID-19/SARS-CoV-2 infection diagnosis, thereby allowing affordable fast and sensitive COVID-19/SARS-CoV-2 laboratory testing.

## Data Availability

No relevant data is linked to the study.

## Acknowledgements

We deeply acknowledge the University Hospital Nuestra Señora de Candelaria board of directors and the executive team for their strong support and assistance in accessing diverse resources used in the study.

## Funding sources

This study was supported by Cabildo Insular de Tenerife (CGIEU0000219140); the agreement with Instituto Tecnológico y de EnergÍas Renovables (ITER) to strengthen scientific and technological education, training, research, development and innovation in Genomics, Personalized Medicine and Biotechnology (OA17/008); Ministerio de Innovación y Ciencia (RTI2018-093747-B-I00 and RTC-2017-6471-1), co-funded by the European Regional Development Funds (ERDF); Lab P2+ facility (UNLL10-3E-783), co-funded by the ERDF and “Fundación CajaCanarias”; and the Spanish HIV/AIDS Research Network (RIS-RETIC, RD16/0025/0011), co-funded by Instituto de Salud Carlos III and by the ERDF.

## Declaration of interests

The authors declare that they have no competing interests.

## Author contributions

JAF, RGM and CF designed the study. JAF, RGM, AIC, DGM, HGC, TMTST and CF participated in data acquisition. JAF, RGM and CF performed the analyses and data interpretation. LC, AVF, RGM and CF wrote the draft of the manuscript. All authors contributed in the critical revision and final approval of the manuscript.

## Notes

### Competing Interest Statement

The authors have declared no competing interest.

